# Body Image Satisfaction and Associated Factors Among Tertiary Students in Harare, Zimbabwe: A Cross-Sectional Study

**DOI:** 10.1101/2025.06.17.25329757

**Authors:** Kaycee R. Gadaga, Ruvimbo Katuka, Nokutenda Charehwa, Prosper V. Sithole, Anotida R. Hove, Shalom R Doyce, Rufaro H. Mushonga, Sidney Muchemwa, Dixon Chibanda, Beatrice K. Shava, Jermaine M. Dambi

## Abstract

**Background:** Body image is a subjective mental representation of an individual’s body appearance, attractiveness, and physical functioning. Lower satisfaction often leads to psychological distress, decreased self-worth and unhealthy habits. This study aimed to determine the prevalence of body image satisfaction and its associated factors among undergraduate university students in Harare, Zimbabwe. Study outcomes will guide the development of bespoke interventions.

**Methods:** Using convenience sampling, this cross-sectional study recruited 1,136 undergraduate students from five universities in Harare, Zimbabwe. Data were self-reported and collected using the Body Appreciation Scale-2, Patient Health Questionnaire-4, Rosenberg Self-Esteem Scale, and socio-demographic questionnaire. Multivariate binary logistic regression analysis was conducted to examine factors influencing body image satisfaction. Data were analysed using SPSS (Version 29) at α=0.05.

**Results:** The prevalence of body image satisfaction was 57.5%. These factors were significantly associated with lower odds of body image satisfaction; being in a relationship [AOR=0.64; (95%CI: 0.47-0.87), p=0.004], smoking [AOR 0.56; (95%CI: 0.33-0.92), p=0.023], engaging in self-body criticism [AOR = 0.31; (95%CI: 0.21-0.47), p< .001], being at risk of anxiety or depression [AOR = 0.72, 95%CI: 0.54-0.95), p=.018], and using diet or exercise as a coping strategy [AOR = 0.71; (95%CI: 0.52-0.96), p=0.027]. However, engaging in daily physical activity [AOR = 2.07; (95%CI: 1.33-3.24),p<0.001], lower daily social media usage [AOR = 1.37; (95%CI: 1.11-1.84), p=.038], a strong sense of belonging and acceptance within university community [AOR = 5.31, (95%CI: 1.49-18.90),p=0.10] and high self-esteem [AOR=3.18; 95%CI: 2.39-4.24),p<.001] were associated with higher odds of body image satisfaction.

**Conclusions:** These findings suggest that interventions aimed at improving body image satisfaction among university students should prioritise psychological well-being, healthy lifestyle behaviours, and a supportive campus environment. Addressing social influences such as self-body criticism and social media engagement may also contribute to more positive body image outcomes in this population.

## Background

Body image is a subjective mental representation of an individual’s body appearance, attractiveness, physical health and functioning [1]. Consequently, body image satisfaction, which is the extent to which an individual is content with their physical appearance, is a multifactorial concept that comprises perceptual, attitudinal and behavioural components [2]. Globally, body image satisfaction is an evolving, dynamic global mental health issue, affecting different contexts and demographics, especially young adults such as university students [3]. Young adults invariably experience different perceptions of their body image, leading to varying levels of body image satisfaction across different settings [4]. These variations are primarily shaped by cultural, geographical and socioeconomic factors, resulting in notable disparities in body image satisfaction across contexts [5]. For instance, a systematic review has yielded variable body image satisfaction prevalence rates ranging from 18-56.6% [6]. Body image satisfaction is a complex concept that includes cognitive, emotional, physical, social, and cultural aspects. Mental health functioning has been identified as an important predictor and outcome of body image satisfaction [7]. For instance, higher body satisfaction enhances mental well-being (e.g. increased self-esteem and self-efficacy); in turn, optimal mental well-being fosters a more positive body image [4]. Inversely, poor body image satisfaction is associated with negative psychological constructs such as low self-esteem, depression, anxiety, substance misuse, eating disorders and stress, while pre-existing mental health challenges can further reinforce negative body perceptions [8]. For example, in an Australian longitudinal study (N=2982), young adults expressing body dissatisfaction exhibited a 10% increase in the rate of binge drinking and tobacco smoking [9]. Systematic reviews further underscore the pervasive impact of body image dissatisfaction, linking it to a broad range of negative outcomes such as psychological distress, disordered eating behaviours, poor sleep quality and diminished academic performance, particularly among university students [10]. Body image satisfaction is also influenced by a variety of factors, including social media usage, peer and familial influence, and socio-cultural dynamics [11]. In the contemporary digital age, plagued with numerous social media platforms, the dynamics surrounding body image concerns have experienced a significant transformation [12]. For instance, constant exposure to carefully curated and frequently digitally altered images of the human body promotes a culture of constant comparison, contributing to widespread body dissatisfaction and psychological distress [13]. Moreover, lifestyle choices such as regular physical activity are correlated with enhanced mental health, while sedentary habits can adversely affect body image satisfaction and mental well-being, thereby perpetuating a detrimental cycle [14]. Consequently, there is an exponential increase in epidemiological studies globally, including bespoke body image interventions [10,15]. However, most of these studies originate from high-income countries, leaving a significant research gap in low- and middle-income countries, such as Zimbabwe, where body image issues remain hugely underexplored. This disparity in the evidence base is concerning, given the emerging global pandemic of body image dissatisfaction and the context-specific/sociocultural conceptualisation of body image satisfaction. To bridge this gap, we sought to assess the prevalence of body image satisfaction and associated factors, among tertiary students in Harare, Zimbabwe. Findings from our study will offer valuable insights into socio-cultural determinants of body image satisfaction, including gathering epidemiological data to inform the development and evaluation of bespoke interventions.

## Materials and Methods

### Study design and setting

This cross-sectional study was conducted between 14 February 2025 and 10 March 2025. Participants were recruited from five tertiary institutions, namely Catholic University in Zimbabwe, Harare Institute of Technology, Harare Polytechnic, University of Zimbabwe and Women’s University in Africa. All the tertiary institutions are in Harare, the capital city of Zimbabwe. Further, the five institutions have a combined enrolment of 34,557 students, approximating 29.7% of tertiary students in Zimbabwe; this increases the study’s external validity.

### Participants

The study population consisted of tertiary students. Students were recruited through convenience sampling, and to be recruited in the study, students had to be 18 years or above, registered as an undergraduate student at any of the five tertiary institutions and available on the day of data collection. Students were excluded from the study only if they were unwilling to provide written consent or were not interested in participating in the study.

### Sample size calculations

Sample size calculations were based on findings from a previous study conducted in Nigeria, where 17.7% (p∘=0.18) of undergraduate students reported high body image satisfaction [16]. Given the contextual similarities, we assumed a comparable but slightly lower expected prevalence estimate of 15% among Zimbabwean students. Based on these parameters, the minimum sample size was 1136 at α = 0.05 and β = 0.80. Sample size calculations were performed using STATISTICA (Version 14).

### Data collection procedures

We first sought and received written permission from the five tertiary institutions to conduct the study. Afterwards, we received ethical approval from the Joint Parirenyatwa Hospital and College of Health Sciences Research Ethics Committee (Ref: JREC/128/25) before commencing data collection. Researchers approached students in public areas such as lecture rooms, benches around the campus or common rooms. Permission was also sought from lecturers to address students in their classes as groups. When students were approached, the study’s purpose was fully explained, and researchers ensured participants understood key ethical principles, including consent, privacy, autonomy and risks. Interested participants signed a consent form before participating in the study. Data were self-completed; participants completed the questionnaires programmed on KoboCollect, using Samsung tablets provided by the research team. The researchers remained nearby to clarify questions or read them out loud if necessary. Upon completion, participants received snacks as a small incentive for participating in the study, and not contingent upon responses.

### Outcome measures

#### Body Appreciation Scale-2

The Body Appreciation Scale-2 (BAS-2) is a 10-item tool to assess body appreciation and positive body image with items rated on a 5-point Likert scale ranging from 1= Never to 5 = Always [17]. Scores are calculated by summing responses of all items and range from 10 to 50, with a higher score indicating a higher body appreciation [17]. The BAS-2 is an extensively applied body image satisfaction scale with robust psychometric performance, including in college and community populations [18,19].

#### Patient Health Questionnaire (PHQ-4)

The four-item Patient Health Questionnaire 4 (PHQ-4) is an ultra-brief self-report depression and anxiety screening tool [20]. Participants rate at which they experience anxiety/depression symptoms on a 4-point Likert scale ranging from 0 (‘not at all’) to 3 (‘nearly every day”), resulting in a total score between 0 and 12, where a higher score denotes a greater likelihood of depression and or anxiety [20]. The PHQ-4 has robust psychometric performance and has been validated in college/university students [20,21].

#### Rosenberg Self-Esteem Scale (RSES)

The Rosenberg Self-Esteem Scale (RSES) is a 10-item questionnaire that measures overall self-worth or self-acceptance [22]. The scale consists of five positively worded statements (e.g., ‘‘I feel that I have a number of good qualities’’) and five negatively worded statements (e.g., ‘‘I feel I do not have much to be proud of’’) with each item rated on a 4-point Likert scale from Strongly Agree (3) to Strongly Disagree (0) [22]. Negatively worded items are reverse-scored, and the scores of all 10 items are summed to get a total score ranging from 0 to 30, with a higher score indicating higher self-esteem. The RSES has excellent validity and reliability and has been validated in university students [22,23].

## Ethical Statement

Written approvals were granted from the study sites/institutions, after which ethical approval was obtained from the Joint Parirenyatwa Hospital and College of Health Sciences Research Ethics Committee (JREC/128/25).

## Data Analysis

Descriptive statistics such as frequencies, percentages and means were computed to describe participant characteristics, prevalence and standardised outcomes. Binary logistic regression was utilised to assess factors related to body image satisfaction. Due to the absence of established cut-off points for the Body Appreciation Scale-2 (BAS-2), the primary outcome, participants were classified based on their mean scores. Participants with scores below the population mean were classified as having low body image satisfaction. This approach was utilised to determine the prevalence of body image satisfaction in this study. Crude odd ratios were calculated, and all variables yielding values p ≤0.20 were fed into the multivariate binary logistic model. Tests were conducted using SPSS Version 30, with the significance level set at α = 0.05.

## Results

### Participant characteristics

Most of the participants were females (51.9%; n=590), single (53.6%; n=609), non-smokers (92.1%; n=1046), non-alcohol drinkers (71.3%; n=810), and did not use drugs (95.8%; n=1088). Nearly all participants (94.7%; n=1076) used social media, mainly WhatsApp (82.7%; n=890), with most spending 2-5 hours daily online (46.0%; n=495). Daily physical activity decreased slightly from 17.1% (n=194) before university to 16.6% (n=181) after enrolment. While 23.6% (n=268) experienced significant weight gain, 24.5% (n=278) reported significant weight loss. Some participants faced body image criticism from family (17.7%; n=201), and friends (18.5%; n=210), while 29.5% (n=335) engaged in self-body criticism. Exercise or diet (28%; n=318), diverting thoughts of body image criticism (29.5%; n=335) and use of positive self-talk/affirmations (28.0%; n=318) were the most used coping mechanisms for body criticism. Last, nearly half (47.2%; n=536) felt very accepted by university peers – See Table 1.

**Table 1:** Participant sociodemographic characteristics (n=1136)

| Variable | Attributes | Frequency, n (%) |
| --- | --- | --- |
| Sex | Male | 546 [48.1] |
|  | Female | 590 [51.9] |
| Relationship status | Single | 609 [53.6] |
|  | Situation-ship | 104 [9.2] |
|  | In a relationship | 361 [31.8] |
|  | Other | 62 [5.5] |
| Smoking Status | No | 1046 [92.1] |
|  | Yes | 90 [7.9] |
| Alcohol Status | No | 810 [71.3] |
|  | Yes | 326 [28.7] |
| Drug and substance status | No | 1088 [95.8] |
|  | Yes | 48 [4.2] |
| Social media use | No | 60 [5.3] |
|  | Yes | 1076 [94.7] |
| Used social media platforms | WhatsApp | 890 [82.7] |
|  | Facebook | 534 [49.6] |
|  | Instagram | 488 [45.4] |
|  | Twitter (X) | 213 [19.8] |
|  | Pinterest | 161 [15.0] |
|  | YouTube | 439 [40.8] |
|  | Tik Tok | 510 [47.4] |
|  | Other | 122 [11.3] |
| Social media frequency | Less than an hour | 134 [12.5] |
|  | 2-5 hours | 495 [46.0] |
|  | More than 5 hours | 447 [41.5] |
| Frequency of physical activity engagement before starting university | Daily | 194 [17.1] |
|  | Several times a week | 279 [24.6] |
|  | Once a week | 145 [12.8] |
|  | A few times a month | 208 [18.3] |
|  | Rarely/never | 310 [27.3] |
| Current physical activity engagement | Daily | 189 [16.6] |
|  | Several times a week | 228 [20.1] |
|  | Once a week | 143 [12.6] |
|  | A few times a month | 228 [20.1] |
|  | Rarely/never | 348 [30.6] |
| Weight Changes | Significant weight gain | 268 [23.6] |
|  | Significant weight loss | 278 [24.5] |
|  | Both weight gain and loss | 298 [26.2] |
|  | No significant changes | 292 [25.7] |
| Body criticism from family | No | 935 [82.3] |
|  | Yes | 201 [17.7] |
| Body criticism from friends | No | 926 [81.5] |
|  | Yes | 210 [18.5] |
| Self-body criticism | No | 954 [84.0] |
|  | Yes | 182 [16.0] |
| Coping strategies | Avoid thinking about it or distract yourself | 335 [29.5] |
|  | Talk to someone | 138 [12.1] |
|  | Exercise or change diet | 317 [27.9] |
|  | Compare yourself to others in person or on social media | 62 [5.5] |
|  | Use positive self-talk or affirmations | 318 [28.0] |
|  | Other | 190 [16.7] |
| Acceptance at university | Very accepted | 536 [47.2] |
|  | Somewhat accepted | 199 [17.5] |
|  | Neutral | 344 [30.3] |
|  | Somewhat unaccepted | 40 [3.5] |
|  | Very unaccepted | 17 [1.5] |

**Table 2:**
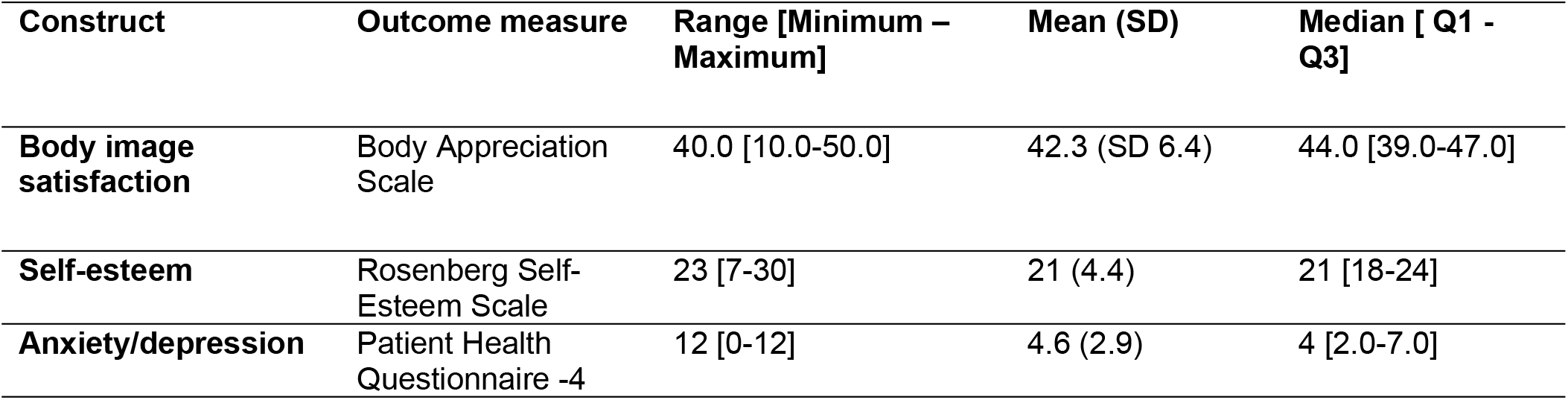
Summative indices.

### Body image, self-esteem and anxiety/depression summative indices

Table 2 shows the study outcome summative indices. The mean (SD) BAS-2 score was 42.3 (SD), giving a 57.5% prevalence of body image satisfaction. Most participants reported being comfortable with their bodies (79.5%; n=903) and respecting them (83.5%; n=980) - See S1 Table. The mean self-esteem score on the Rosenberg self-esteem scale was 21.4 (SD 4.4), with most participants endorsing that they had several good qualities (91.8%; n=1043) and that they could do things as well as others, 90.3% (n=1026) - See S2 Table. The mean PHQ-4 was 4.6 (SD 2.9), with most participants reporting not being able to stop or control worrying (33.8%; n=384) and having little interest or pleasure in doing things (36.8%; n=418) - See S3 Table.

### Factors associated with body image

The S4 Table shows the crude odds ratios of factors associated with body image satisfaction, with Table 3 showing the adjusted analysis. Compared to non-smokers, participants who smoked were 44.5% less likely to have body image satisfaction, AOR=0.56 (95% CI: 0.33 - 0.92), p=0.023. Further, compared to those who were single, participants in relationships were 36.4% less likely to have body image satisfaction, AOR=0.64 (95% CI: 0.47-0.87), p=0.004. Also, participants who engaged in physical activity daily were 2.1 times more likely to have body image satisfaction than those who rarely/never engaged in physical activity, AOR=2.07 (95% CI: 1.33-3.3), p=0.001. Compared to those who spent more than 5 hours on social media, participants who spent 2-5 hours on social media were 1.4 times more likely to have body image satisfaction, AOR=1.37 (95% CI: 1.02-1.84), p=0.038. Furthermore, compared to those who did not criticise themselves, participants who criticised themselves were 68.6% less likely to have body image satisfaction, AOR= 0.31 (95% CI: 0.21-0.47), p=0.001. Participants who felt socially accepted were 5.3 times more likely to have body image satisfaction compared to those who were not socially accepted, AOR=5.31 (95% CI: 1.49-18.90), p=0.010. Compared to those who did not use exercise or dietary changes as coping strategies, participants who exercised or modified their diet were 29.5% less likely to have body image satisfaction, AOR=0.71 (95% CI: 0.52-0.96), p=0.027. Compared to those without anxiety/depression risk, participants who are at risk of having anxiety/depression were 28.5% less likely to have body image satisfaction, AOR=0.72 (95% CI: 0.54-0.95), p=0.018. Last, compared to those with high self-esteem, participants with low self-esteem were 3.2 times less likely to have body image satisfaction, AOR=3.18 (95% CI: 2.39-4.24), p=0.001.

**Table 3:** Factors associated with body image satisfaction.

| Variable | Attributes | Adjusted OR [95% CI] | p-value |
| --- | --- | --- | --- |
| Sex | Male<br>Female (ref) | 0.773 [0.563: 1.060] | 0.110 |
| Relationship status | Situation-ship<br>In a relationship<br>Other<br>Single (ref) | 0.677 [0.413: 1.110]<br>0.636 [0.467: 0.865]<br>1.216 [0.628: 2.355] | 0.122<br>**0.004**<br>0.561 |
| Smoking Status | No<br>Yes (ref) | 0.555 [0.333: 0.924] | **0.023** |
| Social media frequency | Less than an hour<br>2-5 hours<br>More than 5 hours (ref) | 1.570 [0.977: 2.522]<br>1.367 [1.017:1.838] | 0.062<br>**0.038** |
| Physical activity engagement before university | Daily<br>Several times a week<br>Once a week<br>A few times a month<br>Rarely/never (ref) | 2.073 [1.327: 3.239]<br>1.173[0.798: 1.726]<br>1.264 [0.796: 2.008]<br>0.718 [0.479: 1.076] | **<0.001**<br>0.417<br>0.321<br>0.108 |
| Self-body criticism | No<br>Yes (ref) | 0.314 [0.213: 0.465] | **<0.001** |
| Coping strategies | Talk to someone<br>Exercise or change diet | 0.533 [0.570: 1.338]<br>0.705 [0.517: 0.962] | 0.533<br>**0.027** |
| Sense of belonging and acceptance within the university community | Very accepted<br>Somewhat accepted<br>Neutral<br>Somewhat unaccepted<br>Very unaccepted (ref) | 5.312 [1.493: 18.903]<br>1.688 [0.464: 6.140]<br>2.502 [0.700: 8.937]<br>1.245 [0.282:5,508] | **0.010**<br>0.427<br>0.158<br>0.773 |
| PHQ-4 | At risk of having anxiety/depression<br>Anyone without anxiety/depression (ref) | 0.715 [0.540: 0.945] | **0.018** |
| RSES | At risk of having low self-esteem<br>Anyone with high self-esteem (ref) | 3.179 [2.386: 4.236] | **<0.001** |

## Discussion

### Overall synthesis

We sought to assess the prevalence of body image satisfaction and its associated factors among undergraduate students in Harare, Zimbabwe. The prevalence of body image satisfaction in this study population was 57.5%. Being in a relationship, smoking, engaging in self-body criticism, poor mental health, sedentary behaviour, high social media usage, and a low sense of belonging and acceptance within the university community were associated with lower body image satisfaction.

### Prevalence of body image satisfaction

The comparatively high level of body image satisfaction in this study population is comparable to previous studies in the Sub-Saharan region [24,25] and is partly attributed to cultural perceptions of body size and traditional beauty ideals. For example, in many Zimbabwean communities, a fuller body is often associated with health, wealth and social stability, which contrasts with the thinness ideal promoted by Western societies [7]. The culturally rooted perspectives may foster a more accepting attitude towards diverse body types, thereby enhancing body image satisfaction [7]. Nevertheless, our findings diverge from those of a cross-sectional survey among Kenyan college students (N=156) that yielded a positive body image prevalence of 72% [24]. The Kenyan study found that cultural acceptance of body diversity and peer influence played a significant role in fostering positive body image perceptions among students [24]. However, the study’s use of a single institution in Kenya, coupled with a lower sample size, may have led to an overestimation of positive body image, possibly explaining the differences from the more varied sample in our study. In contrast, a cross-sectional study evaluating body image perceptions among Egyptian university students (N=400) yielded a prevalence of 30.75% [26]; this is lower than our study. The Egyptian study attributed the low prevalence of body satisfaction to a strong desire for weight loss, especially among female students, driven by internalised thinness ideals and the belief that men are more attracted to slimmer female bodies; a perception reinforced by cultural and media portrayals of ideal female attractiveness [26]. Collectively, our study further supports the need for context-specific epidemiological data on body image satisfaction, given variations in cultural influences. Such data are essential for the development of bespoke interventions.

### Factors associated with body image satisfaction

#### Self-esteem

In this study, students with low self-esteem were significantly more likely to have lower body satisfaction [AOR=3.18; (95% CI; 2.39-4.24), p < 0.01]. In Zimbabwe, dominant beauty ideals are also prevalent. For instance, there is a cultural emphasis on curvaceous bodies for females, particularly large hips and buttocks; individuals without these physical traits may experience diminished self-worth and self-esteem [27]. Low self-esteem makes individuals vulnerable to internalising societal beauty standards [28], and such individuals may be less likely to seek or receive support, making them more prone to adopt negative body perceptions without challenge, creating a vicious cycle [29]. Our results are consistent with findings from other cultural contexts [26,30,31]. For instance, a study among Egyptian university students (N=400), found that students with lower self-esteem were more likely to have negative body image perceptions (r=0.30, p<0.001), suggesting that social and cultural pressures can strongly influence body image and self-worth among university students [32]. The interplay between external pressures and internal self-worth aligns with the body image resilience theory, which posits that individuals with higher self-esteem are better equipped to resist negative societal pressures and maintain a more positive body image [17]. This theory supports our findings, suggesting that students with lower self-esteem may lack the psychological resources necessary to challenge unrealistic beauty standards, filter out negative influences and maintain body acceptance [33]. Body image satisfaction interventions should have self-esteem enhancement strategies as a core active ingredient.

#### Anxiety and depression

Our study indicates that students at risk of anxiety or depression were significantly less likely to have body image satisfaction [AOR=0.715, (95% CI; 0.540-0.945), p=0.018]. Elsewhere, a Kenyan study among university students (N=620) reported that those experiencing depressive symptoms (measured using PHQ-4) had a 20% lower likelihood of body image satisfaction [AOR=0.802, (95% CI: 0.640-0.965), p=0.043], which is consistent with our findings [34]. Individuals with anxiety and depression often experience negative self-perception, which may lead them to view their bodies more critically and with less appreciation [34]. Additionally, heightened social comparison tendencies common in anxiety may cause individuals to judge their appearances more harshly when comparing themselves to others, contributing to lower body image satisfaction [34]. Elsewhere, a Pakistan study on university students (N=250) found a positive correlation between body image concerns and psychological distress (r=0.63, p<0.01), indicating that higher body image concerns were associated with increased psychological distress [35]. Similarly, a cross-sectional study among Chinese college students (N=150) also demonstrated a negative correlation between body satisfaction and depression and anxiety [r = 0.322, p = 0.001], indicating that lower body dissatisfaction predicts higher levels of mental distress [36]. There is a stern need to address the mental health impacts of body image satisfaction, given the bidirectional relationship between the two constructs.

#### Physical activity

In this study, undergraduate students who engaged in daily physical activity were 2.1 times more likely to have body image satisfaction than those who rarely or never participated in physical activity, AOR=2.07 [(95% CI: 1.33-3.24), p = 0.001]. Engaging in regular physical activity is not only advantageous for physical health but also significantly influences self-perception and psychological well-being. It encourages individuals to appreciate their bodies for their strength and functionality rather than merely appearance, while also enhancing mental well-being by reducing stress and anxiety [37– 39]. According to the Exercise and Self-Esteem Model, regular physical activity engagement enhances physical self-perception and self-worth, which in turn improve global self-esteem and body image [40]. For instance, systematic reviews found that university students who maintained regular physical activity, particularly aerobic and resistance training, reported higher body image satisfaction, increased physical self-worth and lower levels of anxiety [37,41]. Collectively, it’s essential to implement targeted interventions to increase physical activity in university students, with infused body appreciation components.

#### Relationship Status

This study observed that participants in romantic relationships were 36.4% less likely to report body image satisfaction than single participants [AOR = 0.64; 95%CI: (0.47 – 0.87), p=0.004]. The experience of low body image satisfaction for students in relationships may possibly be due to increased self-scrutiny, partner comparison, or fear of not meeting perceived partner expectations, and this is reported elsewhere [42]. Similarly, in a cross-sectional study survey conducted across 10 U.S. universities (N=12,176), they observed that students in relationships were more likely to report body dissatisfaction, especially when partner support for appearance is low, and being in a romantic relationship was associated with increased investment in physical appearance [43,44]. In the US study, individuals in romantic relationships experienced greater sensitivity to appearance-related feedback and increased body monitoring, which can undermine body image satisfaction despite the emotional closeness such relationships provide [43]. Our findings challenge the common assumption that being in a romantic relationship serves as a buffer against body dissatisfaction by providing emotional support and increased self-worth [45,46]. This positive association can be understood through the lens of attachment theory by Hazan and Shaver, which posits that securely attached individuals experience greater self-worth and are more likely to view their partners as emotionally supportive [47]. Such affirming relational dynamics can buffer against external appearance pressures, fostering healthier body image perceptions [47]. This contradiction highlights the complexity of relationship dynamics; while some individuals may experience increased appearance-related pressure, others may benefit from affirming, accepting partnerships that promote positive body image [48].

#### Self-body criticism

In our study, participants who engaged in self-body criticism were 68.6% less likely to be satisfied with their body image than those who did not [AOR = 0.31, (95% CI: 0.21-0.47), p=0.001]. Individuals with frequent self-critical thoughts about their physical appearance are more likely to experience negative evaluations of their body, undermining body image satisfaction [49]. The Self-Discrepancy Theory suggests that body dissatisfaction arises when there is a perceived mismatch between one’s actual body and one’s internalised ideal body [50]. Such discrepancies often fuel self-critical thoughts and emotional distress, especially among young adults exposed to idealised body standards [50]. In Zimbabwean universities, limited access to mental health support means students often internalise stress and low self-esteem, which increases vulnerability to self-criticism and lower body image satisfaction [51]. This finding aligns with a longitudinal study conducted in Australia investigating bi-directional links between self-criticism, self-compassion and positive body image among young adults (N=2982) [9]. In the Australian study, evidence of reciprocal relations emerged with self-criticism, in that higher body appreciation predicted decreased self-criticism, in turn predicting higher body appreciation (r= −0.57, p< 0.001) [9]. The authors explained the negative associations stemming from a self-reinforcing cycle where self-criticism undermines body appreciation, which in turn deepens self-critical thoughts [9]. Elsewhere, a cross-sectional study in U.S college students (N=450) also indicated that higher self-objectification was positively correlated to body image dissatisfaction (r=0.41, p<0.001), highlighting that students who engaged in habitual self-monitoring and critical self-evaluation of their bodies were more likely to experience negative body image outcomes [52]. Therefore, there is a need for targeted interventions that also address the impacts of self-criticism/comparisons on body image satisfaction.

#### Social media frequency

In our study, frequent social media users were significantly less likely to report body image satisfaction than infrequent users [AOR = 0.51, (95% CI: 0.40-0.66), p = 0.002]. Our findings reflect a growing global concern that social media platforms, originally designed to foster social connection(s), may instead be fuelling widespread body dissatisfaction [53,54]. Unfortunately, social media frequently presents narrow and often unattainable beauty standards, which predispose users to body image dissatisfaction [55,56]. The intense pursuit of online validation, through likes and comments, further exacerbates body dissatisfaction by influencing how individuals shape their self-image and identity in a world dominated by digital media [57]. Our study findings align with a cross-sectional survey among Nigerian university students (N=542), which showed that increased social media use significantly increased body image dissatisfaction (r=0.58, p < 0.001) [58]. Similarly, a cross-sectional study among Kenyan undergraduate students (N=400) attributed lower body image satisfaction to excessive social media use with resultant increased exposure to Western media promoting unrealistic beauty standards, poor self-esteem and mental health issues [19]. Elsewhere, a cross-sectional study of 1087 university students in Australia found that frequent social networking site use predicted greater internalisation of thin body ideals (β= 0.34, p<0.001) and higher levels of body surveillance (β=0.29, p<0.001) [59], further highlighting the role of social media in reinforcing unattainable beauty standards. Overall, our findings support global evidence that frequent/excessive social media use negatively affects body image satisfaction among university students, hence the need for bespoke and holistic interventions that address both excessive social media usage and body image dissatisfaction.

#### Sense of belonging and acceptance within the university community

Our study found that undergraduate students who felt a sense of belonging and acceptance within the community were 5.3 times more likely to report higher body image satisfaction compared to those who did not, AOR=5.31 [95%CI: (1.49-18.90), p <0.001]. A strong sense of belonging is a well-established protective factor that enhances psychological well-being and mitigates mental health challenges, including body dissatisfaction [60]. When students feel accepted and valued, they are less likely to conform to unrealistic body ideals or engage in harmful social comparisons. This fosters self-acceptance and appreciation of their own appearance [61,62]. Similarly, a cross-sectional study among US university students (N=465) indicated that students who reported high levels of social support, particularly those who felt they could talk to a family member about important matters, or felt loved by a family member or friend, exhibited higher body image satisfaction [63]. The body image satisfaction/social acceptance association aligns with the self-determination theory, which posits that psychological well-being is fostered when individuals’ basic needs for relatedness, autonomy and competence are met [64]. Feeling emotionally supported helps college students manage stress and develop a more positive view of their bodies, while affirming feedback from friends and loved ones enhances self-worth and reduces the impact of unrealistic beauty ideals [65]. It is essential to implement interventions that promote social inclusion, acceptance, and connectedness to safeguard individuals from internalising detrimental appearance ideals and to encourage a more comprehensive appreciation of their bodies. Furthermore, a strong sense of belonging within the university community decreases the likelihood of harmful social comparisons, encouraging greater acceptance of one’s own appearance [62].

#### Coping strategies

In our study, participants who engaged in exercise and dietary changes as coping strategies in response to body dissatisfaction or body image criticism reported higher levels of body image satisfaction. In contrast, those who relied on strategies such as comparing themselves to others on social media, avoidance, distraction, or positive self-talk and affirmations tended to report lower body image satisfaction [AOR= 0.71, 95% CI: (0.52-0.9622), p = 0.001]. Engagement in positive health behaviours like exercise and dietary modification may serve as positive adaptive coping mechanisms, leading to optimal body image satisfaction outcomes [39]. However, not all engagement in exercise and dieting necessarily stems from a desire to enhance appearance, as individuals may adopt these behaviours for intrinsic health-related motivations, such as improving physical fitness, enhancing mental well-being, or managing chronic conditions [66,67]. A cross-sectional study among Swedish university students (N=4,262) provides additional insights into the protective functions of regular exercise on body image satisfaction [68]. Also, the Swedish study points to the use of exercise as a maladaptive stress-coping strategy driven by internalised body ideals and perfectionism [68]. Therefore, while appearance-driven motives cannot be ruled out, particularly in a sociocultural context that idealises certain body types, the association observed in our findings may reflect a complex interplay of both health-focused and appearance-focused physical activity engagement goals [69]. Another cross-sectional study conducted among Nigerian university students (N=865) reports the use of dietary control as a salient strategy for the attainment of the desired body image [70]. The authors suggested that sociocultural pressures and media portrayals of ideal bodies likely influenced students’ reliance on dietary changes as a means of improving appearance [70]. Therefore, it is essential to include healthy lifestyle counselling components that foster a positive body image holistically, as exercise and dietary modifications can contribute to a healthy lifestyle; they can also be equally used as maladaptive coping mechanisms for body dissatisfaction.

#### Smoking

Our study found that participants who smoked were 44.5% less likely to have body image satisfaction [AOR=0.56; 95% CI: (0.33-0.92), p=0.023], supporting existing evidence linking smoking to body image concerns and its role as a maladaptive coping strategy for psychological distress[71,72]. Comparable findings have been reported among Ethiopian university students (N=1029), where smoking was associated with increased odds of body dissatisfaction [AOR = 1.63; 95%CI: (1.03–2.58), p<.001] [71], and in a Swedish cohort of 30,245 young adults, where current smokers were more likely to report negative body image [AOR = 1.60; 95% CI: (1.38–1.87), p < 0.001] [72]. These associations likely reflect shared psychological and behavioural risk factors, including emotional dysregulation and weight control motives [71,72]. In Zimbabwe and similar contexts, smoking is highly stigmatised; the social exclusion may reinforce internalised shame and poor self-image [73,74]. Smoking also produces visible physical effects, such as skin ageing, dental discolouration, and weight fluctuations, which may exacerbate dissatisfaction, particularly in appearance-conscious university settings [75]. Additionally, recognition of the negative impacts of smoking can create internal conflict as individuals strive to align with health and aesthetic ideals, which can influence their self-perception [73,74]. Collectively, our study reaffirms the cyclical relationship wherein body dissatisfaction may contribute to smoking initiation, while continued smoking reinforces negative self-perception through physical effects and psychosocial stressors, hence the need for holistic targeted interventions.

## Conclusion

This study assessed body image satisfaction among 1,136 undergraduates from five tertiary institutions in Harare, Zimbabwe. It found that 57.5% of students were satisfied with their body image. Factors negatively impacting satisfaction included smoking, being in a relationship, heavy social media use, self-body criticism, low self-esteem, anxiety and depression. On the contrary, daily physical activity, social acceptance and healthy coping strategies such as exercise were positively associated with body image satisfaction. These findings highlight the significant impact of lifestyle, social environment, and mental health on body image, underscoring the need for targeted interventions within tertiary institutions to promote body positivity and psychological well-being.

## Data Availability

The datasets used and/or analysed during the current study are available from the corresponding author on reasonable request. The datasets will be made available on online repositories once all manuscripts related to the study have been published online.

## Strengths and limitations

This is one of the few large-scale studies exploring the impact of body image satisfaction in tertiary students in Sub-Saharan Africa. The recruitment of a large sample across multiple sites enhances the study’s external validity. Although the use of a cross-sectional study design cannot infer causality, this study provides bespoke epidemiological data on the burden and determinants of body image satisfaction, which will inform the development of context-specific interventions. However, the use of convenience sampling may have introduced selection bias; randomisation was not possible owing to a limited budget. Also, data were self-reported, and there is a risk of social desirability bias. However, the study explores latent constructs, and the lack of objective indicators is uncommon in global mental health. Last, the Body Appreciation Scale (the study’s primary outcome measure) and the Rosenberg Self-Esteem have not been validated in the research setting; this may have introduced measurement bias. However, these outcome measures are widely used in psychological and public health research, with demonstrated psychometric robustness, enabling comparability of our study outcomes with other studies. Nevertheless, future studies are required for the formal trans-cultural adaptation and validation of these outcome measures.

## Declarations

## Ethics approval and Consent to participate

Ethical approval for the study was granted by the University of Zimbabwe Directorate and the Joint Research and Ethics Committee for the University of Zimbabwe, Faculty of Medicine and Health Science & Parirenyatwa Group of Hospitals (Ref: JREC/128/25). All methods were carried out in accordance with the Joint Research and Ethics Committee guidelines and regulations, which conform to The Declaration of Helsinki. Participants were treated as autonomous agents and were requested to sign written consent before participation. Written informed consent was obtained from all subjects prior to participation in this study. Randomised participant ID numbers were used to preserve confidentiality. Data and signed informed consent forms were stored securely, only the researchers had access to the information gathered, and participants could voluntarily withdraw from the study without any consequences.

## Authors contributions

- Kaycee R. Gadaga (KRG) developed the concept and design of the study, collected the data and drafted the first version of the manuscript.
- Ruvimbo Katuka (RK) developed the concept and design of the study, collected the data and jointly drafted the first version of the manuscript.
- Nokutenda Charehwa (NC) developed the concept and design of the study, collected the data and contributed to the editing of the second through fifth versions of the manuscript.
- Prosper V. Sithole (PSV) developed the concept and design of the study, collected the data and contributed to the editing of the second through fifth versions of the manuscript.
- Anotida R. Hove (ARH) co-supervised the project, critically appraised/peer-reviewed and made substantive contributions to the second to fifth versions of the manuscript in preparation for submission to the journal.
- Shalom R. Doyce critically appraised/peer-reviewed and made substantive contributions to the second to fifth versions of the manuscript in preparation for submission to the journal.
- Rufaro Hamish Mushonga critically appraised/peer-reviewed and made substantive contributions to the second to fifth versions of the manuscript in preparation for submission to the journal.
- Sidney Muchemwa (SM) co-supervised the project, critically appraised/peer-reviewed and made substantive contributions to the second to fifth versions of the manuscript in preparation for submission to the journal.
- Dixon Chibanda provided overall mentorship and critically appraised/peer-reviewed and made substantive contributions to the second to fifth versions of the manuscript in preparation for submission to the journal.
- Beatrice K Shava (BKS) co-supervised the project, critically appraised/peer-reviewed and made substantive contributions to the second to fifth versions of the manuscript in preparation
- for submission to the journal. BKS prepared all prerequisite processes for article submission, submitted the manuscript and is the corresponding author.
- Jermaine M. Dambi (JMD) - developed the concept and design of the study, was the study project main supervisor, conducted the data analysis and statistical interpretation, revised and made substantive contributions to the first to fifth versions of the manuscript
- All authors read and approved the final manuscript.

## Acknowledgements

We would want to acknowledge participants for their invaluable participation, especially. The data were collected as part of KRG, RK, NC & PVS’s undergraduate thesis, which ARH, SM, BKS and JMD supervised.

## Supporting information

**S1 Table: BAS-2 frequencies**

**S2 Table: Rosenberg Self-Esteem Scale Frequencies**

**S3 Table: Patient Health Questionnaire Frequencies**

**S4 Table: Unadjusted odds ratios-body image determinants**

